# Are Emergency Departments becoming inaccessible for vulnerable groups of patients? A retrospective cohort study

**DOI:** 10.64898/2026.10.18.26365831

**Authors:** Jo Mitchell, Simona Hapca

**Affiliations:** Emergency Medicine Consultant, Forth Valley Royal Hospital, Stirling Road FK5 4WR; School of Computing Science and Mathematics, University of Stirling, Stirling FK9 4LA

## Abstract

**Background:** Emergency Department (ED) overcrowding is associated with adverse outcomes including delays to treatment, patients leaving without being seen and increased mortality. Increasingly, evidence suggests that crowding may disproportionately affect those of lower socioeconomic status, worsening health inequalities. This study investigates the relationship between prolonged ED length of stay and vulnerable patient groups choosing not to wait to be seen.

**Methods:** A retrospective cohort study analysed monthly ED attendance data from a large district general hospital between January 2020 and July 2025. Patients who did not wait to be seen (DNW) were assessed for vulnerability based on age, presenting complaint and inferred frailty. Associations were explored using generalised estimation equation model with binary outcome.

**Results:** During the 66-month study period, 317,978 patients attended the ED, of whom 35,403 (11.1%) did not wait to be seen. Of these, just under one third met vulnerability criteria. As the percentage of patients staying in the ED for more than 8-hours increased, so did the proportion of patients leaving the department without being seen. With each 10% increase in LOS>8h percentage, the odds of any patient choosing not to wait increases by 48% (OR=1.48, 95% CI(1.47 – 1.49), p<0.001). Of the three vulnerability groups assessed, paediatric patients, those presenting with mental health complaints and frailty, frail patients were the most affected when lengths of stay in the ED increased and crowding became a problem (OR=1.73, 95% CI(1.72 – 1.74), p<0.001).

**Conclusion:** These findings demonstrate that while all patient groups are more likely to leave without being seen as crowding worsens, elderly and paediatric patient cohorts are significantly more likely to leave without being seen as crowding worsens. This raises genuine concerns about the accessibility of healthcare, in the presence of crowding, in these vulnerable groups.

## Introduction

There is clear evidence in the literature that overcrowding in Emergency Departments (EDs) has adverse effects, including delays to treatment, patients leaving without being seen and increases in mortality^1-4^ . We know that crowding kills^5^, the Royal College of Emergency Medicine has published guidance to assist in the understanding of the causes and effects of crowding and to provide support for health systems^6^. What is becoming evident in the authors practice, however, is that crowding and access to Emergency Care appears to be disproportionally affecting vulnerable groups, specifically, frail, mental health and paediatric patients.

ED’s are a vital point of access into the NHS, especially for those in lower socioeconomic groups^7^. Factors affecting ED attendance are increasingly complex^8^ and poorly understood, despite many researchers seeking to understand this relationship. The number of patients that did not wait (DNW) to be seen have been shown to reflect ED crowding and are commonly used as performance markers in the literature^9^, however, there is no universally acceptable DNW threshold in emergency medicine literature and DNW data is not collected nationally in Scotland. Patients that did not wait are a significant area of concern, with a recent study concluding that patients that DNW were more likely, than patients that were seen and discharged, to re-present to the ED, require admission and die within 30-days^10^.

This study was carried out in a busy district general ED that has an annual attendance figure of around 60,000 patients, and currently has the worst trust-level performance against the 4-hour emergency access standard in NHS Scotland^11^. Despite a sustained drop in 4-hour performance since May 2020, no measures taken have made a significant difference to performance and, perception amongst the clinical team is that the impact of crowding is worse due to an increase in the numbers of patients with length of stays over 8 and 12-hours in the ED^12^. This has obvious implications for staff wellbeing and morale with studies finding statistically significant relationships between higher levels of crowding and increased burnout amongst ED clinicians^9,13^ .

The aim of this study is to investigate the relationship between increased numbers of patients with lengths of stay over 8-hours within the ED and vulnerable patient groups choosing not to wait (DNW) using retrospective analysis.

## Method

For this retrospective cohort study, monthly data of patients from Forth Valley Royal Hospital (FVRH) in Scotland, who did not wait to be seen were collected over a 5-year period, from January 2020 to end June 2025 from Trakcare by Intersystems and accessed through an access request to the NHS FV IT department. FVRH is a busy district general hospital situated within NHS Forth Valley serving approximately 6% of Scotland’s population. FVRH Emergency Department sees around 60,000 patients annually and is the sole Emergency Department within this health board. This period was chosen as it runs through a clear time of drop off in the performance of the 4-hour access standard, not just in this department, but across Scotland as a whole. Patient identifying data were not collected, just patient attendance month, attendance ID number, registered GP, age, presenting complaint and triage information. No exclusions were applied, sample size calculation was not performed as all eligible participants were included in the study. Information was also collected on whether the patient reattended the ED within 7-days of their initial attendance. Patients were reviewed for vulnerabilities by looking at their presenting complaint and triage summary. The number of patients with vulnerabilities were recorded and analysed.

In this cohort study the exposure variable is the length of stay in the ED, with particular focus on lengths of stay in the department of more than 8-hours (LOS>8h). This is not the same as the waiting time to be seen. Patients with a length of stay in the ED of more than 8 and 12 hours (from arrival in ED to exit from ED) are collated by Public Health Scotland and are publicly accessible.

The outcome variable was the number of people that leave without being seen (DNW), which was analysed as a total and in different vulnerability groups.

The vulnerabilities considered were:

1. Whether they were paediatric attenders (under 18-years old at the time of their attendance)
2. Those between 18 and 75-years old presenting with a mental health complaint. These included, but were not limited to, those presenting with overdose, self-harm, self-declared mental health crisis or as a direct result of alcohol or drug intoxication
3. Potentially frail - as no formal frailty scoring occurred at triage, an age of over 75-years was set as a marker for frailty, accepting that many of this patient group are not in fact frail, but allowing this higher age to compensate for the fact that there are many patients under the age of 75 who meet frailty criteria.

All patients recorded as DNW during the study period were included; no cases were excluded due to missing demographic or triage data.

Monthly data regarding number of people with LOS>8hrs and total attendances were accessed publicly through the Public Health Scotland website. Data were analysed in R statistical package “geepack” using a generalised estimation equation model with binomial distribution and autoregressive correlation structure AR(1) (accounting for the temporal autocorrelation), to model the monthly proportion of those who did not wait (Prop.DNW) from the total of people attending the ED, depending on the percentage of patients with a length of stay than 8 hrs within the ED. The analysis was firstly performed for the whole cohort of people attending the ED who did not wait, then for the three vulnerable groups of people, the under 18, the group of 75 years old and above, and the group of people with mental health presentations. The authors verified all code, interpreted the outputs and take full responsibility for the accuracy of the results. There were no missing data, all analyses were unadjusted as we were analysing patient level data.

Patients and public were not involved in the design or running of this study.

## Results

During the 66-months of data collection 317,978 patients attended the ED, of whom 35,403 (11.1%) did not wait to be seen (DNW) The oldest patient who did not wait to be seen was 101, the youngest was just a week old. The average age of patients not waiting to be seen was 34years with a median of 32-years. The records of DNW patients were individually reviewed for vulnerability criteria as defined above (See figure 1). Of the 35,403 patients, 1,388 (3.9%) did not have a registered GP. 31% of patients who did not wait to be seen were assessed as meeting our defined vulnerability criteria (N=11,055). Of these, 74% were paediatric patients (N=8,131), 12% were patients with mental health presentations (N=1,325) and 14% were patients over the age of 75 (N=1,599).

**Figure 1.**
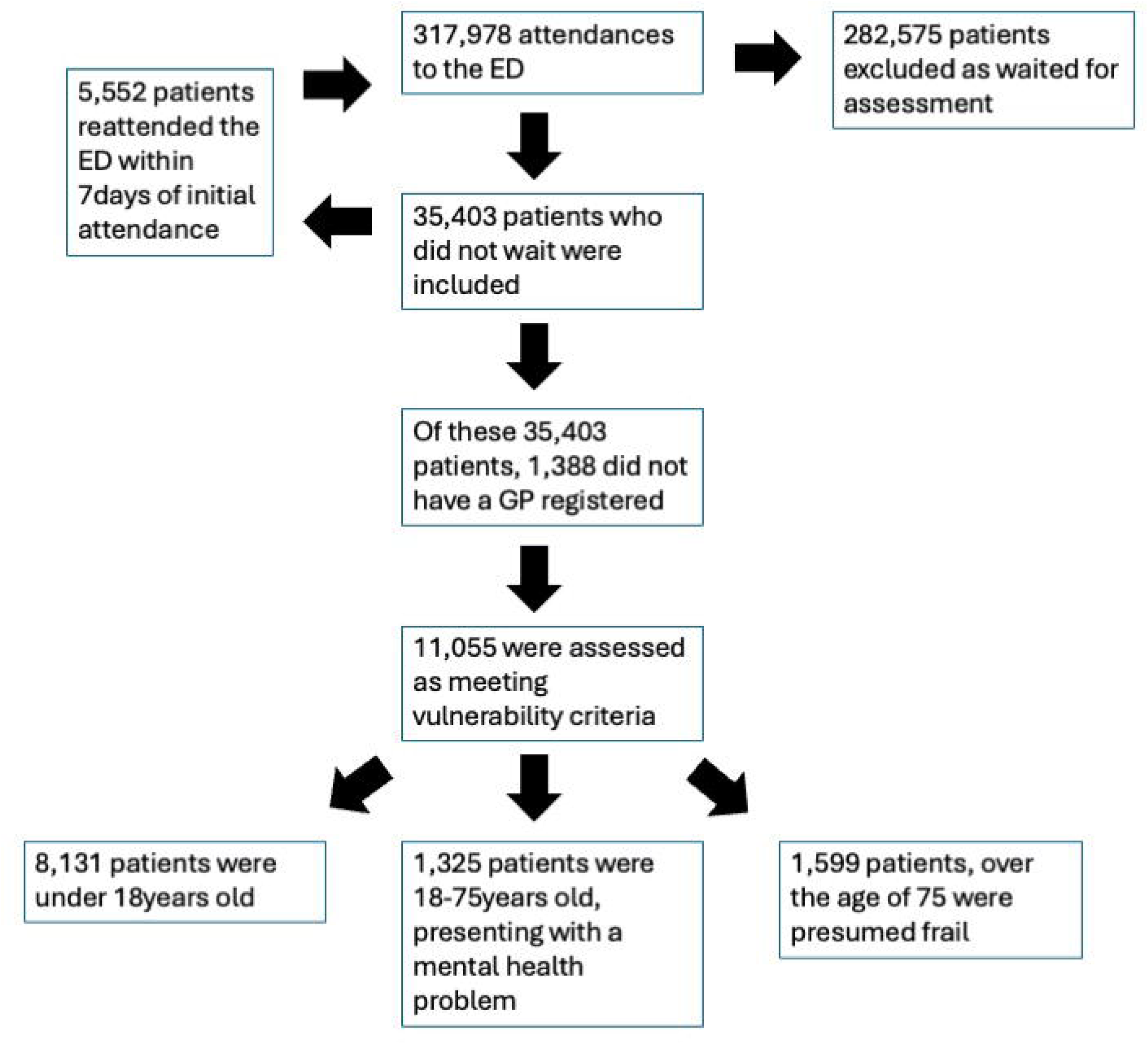
Flow chart showing the flow of patients in the study, along with inclusion and exclusion criteria

Data collected is summarised in Table 1. The average percentage of patients who did not wait to be seen each month was 10.74%, with a minimum number of 1.17% and a maximum figure of 17.34%. Reattendance figures showed an average 7-day return rate of 15.84%, with a minimum of 9.54 % and a maximum of 35.77%. During the same period the average performance with the 4h access standard was 60.52%, with a minimum compliance of 40.2% and a maximum compliance of 97.1% per month. The percentage of patients each month who did not wait (DNW) that were considered vulnerable ranged from 9.54% to 35.77% with an average of 15.84%.

**Table 1.** Summary of data collected for analysis.

| Month | Attendances | LOS>8h (%) | Total DNW | Reattended | Attendances (Numbers) |  |  | DNW (Numbers) |  |  |  |
| --- | --- | --- | --- | --- | --- | --- | --- | --- | --- | --- | --- |
|  |  |  |  |  | <18 | >75 | MH | No GP | <18 | >75 | MH |
| Jan 2020 | 5438 | 1% | 296 | 39 | 879 | 790 | 104 | 10 | 67 | 3 | 7 |
| Feb 2020 | 5016 | 0% | 174 | 22 | 939 | 690 | 129 | 6 | 41 | 5 | 8 |
| Mar 2020 | 4258 | 0% | 156 | 29 | 747 | 659 | 125 | 6 | 30 | 2 | 15 |
| Apr 2020 | 2586 | 0% | 51 | 7 | 287 | 485 | 47 | 4 | 4 | 0 | 4 |
| May 2020 | 3281 | 0% | 58 | 9 | 442 | 530 | 67 | 2 | 9 | 2 | 4 |
| Jun 2020 | 3834 | 0% | 91 | 14 | 508 | 627 | 115 | 4 | 12 | 0 | 4 |
| Jul 2020 | 4155 | 0% | 148 | 23 | 599 | 699 | 169 | 8 | 27 | 5 | 8 |
| Aug 2020 | 4293 | 0% | 218 | 41 | 673 | 741 | 194 | 5 | 49 | 6 | 22 |
| Sep 2020 | 3982 | 0% | 191 | 20 | 594 | 710 | 183 | 7 | 41 | 3 | 12 |
| Oct 2020 | 3808 | 0% | 146 | 37 | 502 | 723 | 163 | 7 | 22 | 11 | 13 |
| Nov 2020 | 3744 | 1% | 123 | 44 | 537 | 622 | 169 | 1 | 14 | 26 | 13 |
| Dec 2020 | 3446 | 0% | 97 | 11 | 449 | 661 | 151 | 0 | 15 | 5 | 10 |
| Jan 2021 | 3517 | 2% | 82 | 14 | 446 | 666 | 178 | 3 | 12 | 2 | 12 |
| Feb 2021 | 3294 | 1% | 116 | 26 | 486 | 567 | 173 | 2 | 14 | 5 | 14 |
| Mar 2021 | 4332 | 1% | 190 | 29 | 856 | 627 | 196 | 2 | 36 | 6 | 14 |
| Apr 2021 | 4746 | 1% | 233 | 44 | 988 | 722 | 183 | 4 | 48 | 3 | 27 |
| May 2021 | 5350 | 1% | 292 | 47 | 1184 | 760 | 182 | 7 | 66 | 6 | 20 |
| Jun 2021 | 5385 | 1% | 483 | 78 | 1184 | 723 | 183 | 10 | 126 | 10 | 25 |
| Jul 2021 | 5268 | 3% | 502 | 78 | 895 | 777 | 209 | 17 | 102 | 17 | 35 |
| Aug 2021 | 5355 | 5% | 561 | 91 | 1035 | 794 | 178 | 27 | 130 | 18 | 21 |
| Sep 2021 | 5373 | 7% | 753 | 124 | 1155 | 776 | 159 | 21 | 195 | 27 | 32 |
| Oct 2021 | 5039 | 14% | 684 | 98 | 1027 | 768 | 159 | 31 | 178 | 23 | 40 |
| Nov 2021 | 4761 | 10% | 492 | 81 | 1055 | 697 | 147 | 8 | 147 | 12 | 26 |
| Dec 2021 | 4537 | 6% | 375 | 44 | 788 | 716 | 141 | 13 | 90 | 13 | 24 |
| Jan 2022 | 4421 | 3% | 278 | 61 | 794 | 676 | 152 | 9 | 71 | 6 | 19 |
| Feb 2022 | 4436 | 3% | 395 | 59 | 843 | 652 | 149 | 8 | 90 | 14 | 29 |
| Mar 2022 | 5134 | 13% | 593 | 94 | 1030 | 815 | 154 | 24 | 162 | 21 | 38 |
| Apr 2022 | 4912 | 10% | 580 | 94 | 899 | 745 | 179 | 36 | 156 | 24 | 39 |
| May 2022 | 5476 | 12% | 696 | 113 | 1128 | 783 | 141 | 27 | 165 | 31 | 26 |
| Jun 2022 | 5321 | 16% | 745 | 100 | 1054 | 771 | 171 | 38 | 179 | 28 | 39 |
| Jul 2022 | 4923 | 19% | 647 | 90 | 869 | 758 | 136 | 18 | 170 | 25 | 28 |
| Aug 2022 | 5023 | 20% | 743 | 107 | 942 | 731 | 149 | 39 | 190 | 28 | 33 |
| Sep 2022 | 4988 | 28% | 864 | 109 | 1010 | 800 | 160 | 27 | 224 | 53 | 48 |
| Oct 2022 | 4911 | 29% | 744 | 98 | 868 | 825 | 143 | 37 | 176 | 37 | 29 |
| Nov 2022 | 4709 | 18% | 499 | 84 | 981 | 682 | 95 | 21 | 138 | 19 | 19 |
| Dec 2022 | 4911 | 26% | 681 | 65 | 1064 | 823 | 73 | 36 | 211 | 32 | 19 |
| Jan 2023 | 4199 | 25% | 380 | 60 | 705 | 707 | 72 | 10 | 83 | 14 | 9 |
| Feb 2023 | 4065 | 20% | 353 | 60 | 815 | 643 | 100 | 14 | 92 | 9 | 16 |
| Mar 2023 | 4930 | 25% | 689 | 84 | 958 | 816 | 80 | 27 | 177 | 27 | 22 |
| Apr 2023 | 4730 | 18% | 526 | 77 | 883 | 731 | 79 | 21 | 114 | 20 | 12 |
| May 2023 | 5381 | 18% | 683 | 111 | 1019 | 773 | 99 | 37 | 154 | 24 | 21 |
| Jun 2023 | 5140 | 17% | 666 | 74 | 1044 | 741 | 76 | 26 | 166 | 30 | 6 |
| Jul 2023 | 5237 | 15% | 620 | 110 | 853 | 752 | 103 | 42 | 132 | 28 | 19 |
| Aug 2023 | 5482 | 16% | 633 | 101 | 1007 | 809 | 115 | 29 | 115 | 35 | 23 |
| Sep 2023 | 5387 | 22% | 768 | 112 | 1062 | 784 | 90 | 39 | 154 | 34 | 20 |
| Oct 2023 | 5165 | 25% | 640 | 99 | 934 | 770 | 81 | 20 | 116 | 23 | 13 |
| Nov 2023 | 5138 | 25% | 693 | 111 | 1020 | 822 | 75 | 26 | 148 | 42 | 19 |
| Dec 2023 | 5109 | 29% | 705 | 98 | 906 | 867 | 76 | 22 | 161 | 38 | 13 |
| Jan 2024 | 4999 | 30% | 650 | 109 | 842 | 798 | 83 | 20 | 146 | 31 | 24 |
| Feb 2024 | 4860 | 30% | 741 | 95 | 881 | 733 | 88 | 44 | 141 | 36 | 9 |
| Mar 2024 | 5345 | 28% | 883 | 131 | 1111 | 826 | 85 | 27 | 248 | 35 | 20 |
| Apr 2024 | 5252 | 25% | 779 | 126 | 951 | 826 | 81 | 25 | 158 | 46 | 24 |
| May 2024 | 5593 | 22% | 749 | 101 | 1070 | 862 | 83 | 20 | 152 | 30 | 21 |
| Jun 2024 | 5358 | 23% | 857 | 147 | 1069 | 787 | 96 | 35 | 186 | 37 | 20 |
| Jul 2024 | 5093 | 18% | 593 | 110 | 815 | 821 | 115 | 23 | 113 | 20 | 24 |
| Aug 2024 | 5327 | 23% | 734 | 132 | 945 | 829 | 86 | 46 | 151 | 36 | 26 |
| Sep 2024 | 5372 | 25% | 858 | 112 | 1001 | 881 | 80 | 27 | 173 | 59 | 16 |
| Oct 2024 | 5335 | 26% | 881 | 127 | 905 | 821 | 88 | 28 | 193 | 55 | 22 |
| Nov 2024 | 5047 | 26% | 781 | 116 | 995 | 791 | 64 | 23 | 183 | 45 | 13 |
| Dec 2024 | 5115 | 30% | 852 | 110 | 987 | 850 | 68 | 27 | 213 | 55 | 16 |
| Jan 2025 | 4967 | 26% | 659 | 102 | 823 | 865 | 57 | 21 | 129 | 35 | 10 |
| Feb 2025 | 4634 | 21% | 588 | 105 | 835 | 729 | 101 | 14 | 132 | 29 | 23 |
| Mar 2025 | 5398 | 22% | 870 | 151 | 1149 | 800 | 98 | 21 | 220 | 52 | 23 |
| Apr 2025 | 5271 | 19% | 679 | 103 | 990 | 870 | 88 | 31 | 143 | 40 | 21 |
| May 2025 | 5665 | 23% | 979 | 142 | 1117 | 904 | 89 | 43 | 245 | 52 | 21 |
| Jun 2025 | 5421 | 23% | 937 | 173 | 988 | 838 | 99 | 31 | 186 | 54 | 23 |

Figure 2 illustrates monthly trends in this ED’s performance indicators over time, allowing direct comparison of their relative movement. A clear inverse relationship is evident between 4-hour standard compliance and the proportion of patients experiencing prolonged waits. As 4-hour compliance steadily declines from 2020 onwards, both LOS>8h and LOS>12h show marked increases, particularly from 2021 onwards. The data highlights how periods of falling compliance align closely with rising long-stay pressures, suggesting worsening system performance rather than short-term fluctuation.

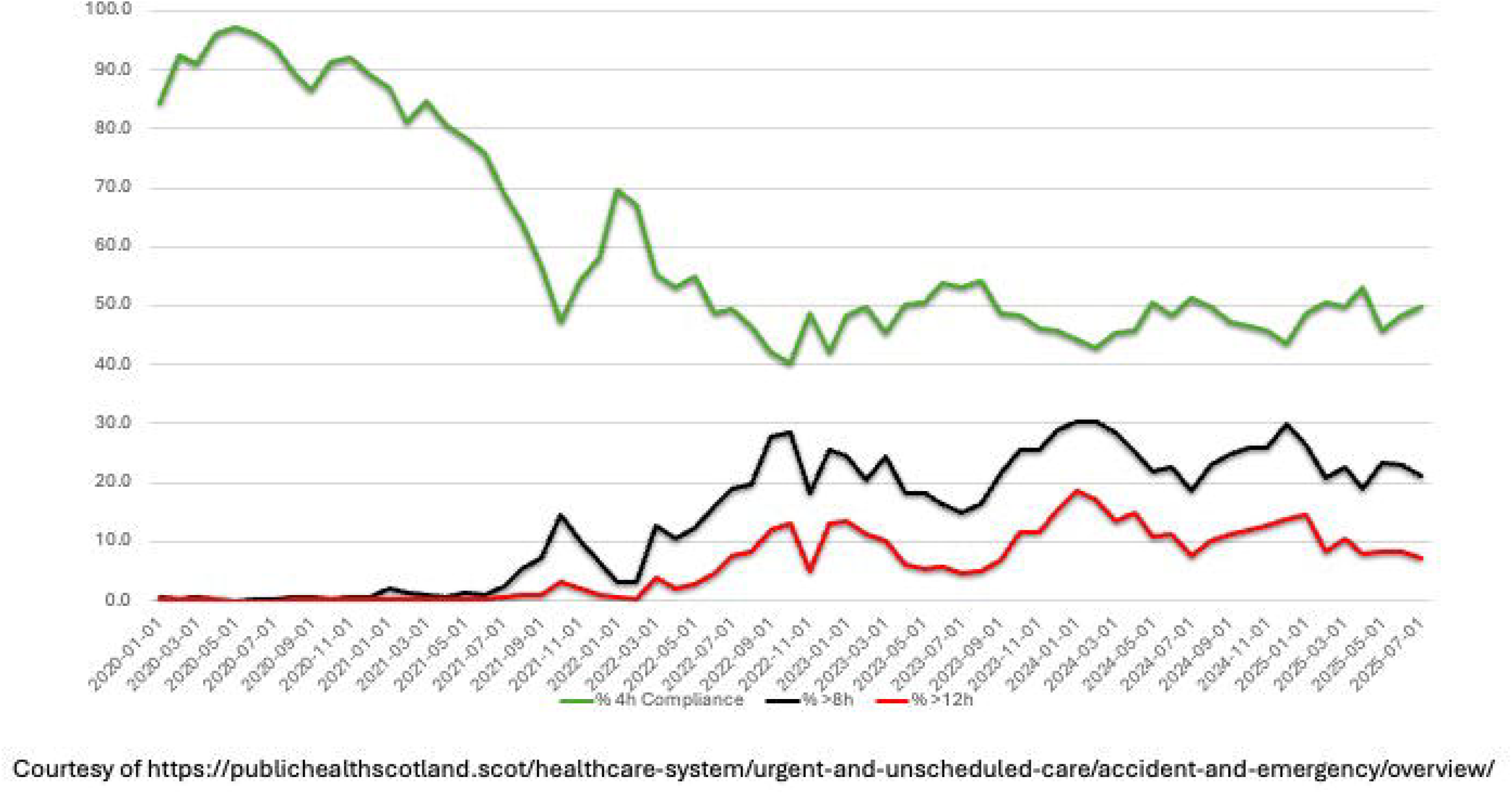

Proportional representation of attendances vs DNW were considered in the ED population as a whole and for each vulnerability group (Figure 3). In early analyses, before proportional representation was considered, patients presenting with mental health conditions appeared to be less likely to leave without being seen as LOS>8h increased. However, you can clearly see from the graphs that from late 2022, numbers of patients presenting to the ED with mental health conditions has reduced. When this group is considered proportionately, the odds of this group of patients leaving without being seen increase similarly to the general population.

Odds ratios between proportion of patients in the ED that did not wait (Figure 4) and length of stay in the ED of more than 8-hours were calculated. In general, with each 10% increase in LOS>8h percentage, the odds of any patient choosing not to wait increases by 48% (OR=1.48, 95% CI(1.47 – 1.49), p<0.001). The odds ratio confirms strong positive associations between the DNW proportion of the individual vulnerable groups and LOS>8h percentage in the ED (p<0.001). Generalised estimation equation model for binomial data, demonstrated that the odds of vulnerable patients that DNW across all groups increased in line with LOS>8h with frail patients 73% more likely to leave without being seen with each 10% increase in LOS>8h (OR=1.73, 95% CI(1.72 – 1.74), p<0.001). Paediatric patients, were 56% more likely to leave without being seen (OR=1.56, 95% CI(1.55 – 1.57), p<0.001), and patients presenting with mental health conditions were 42% more likely to leave without being seen with each 10% increase in LOS>8h (OR=1.42, 95% CI(1.41 – 1.43), p<0.001.

**Figure 3.**
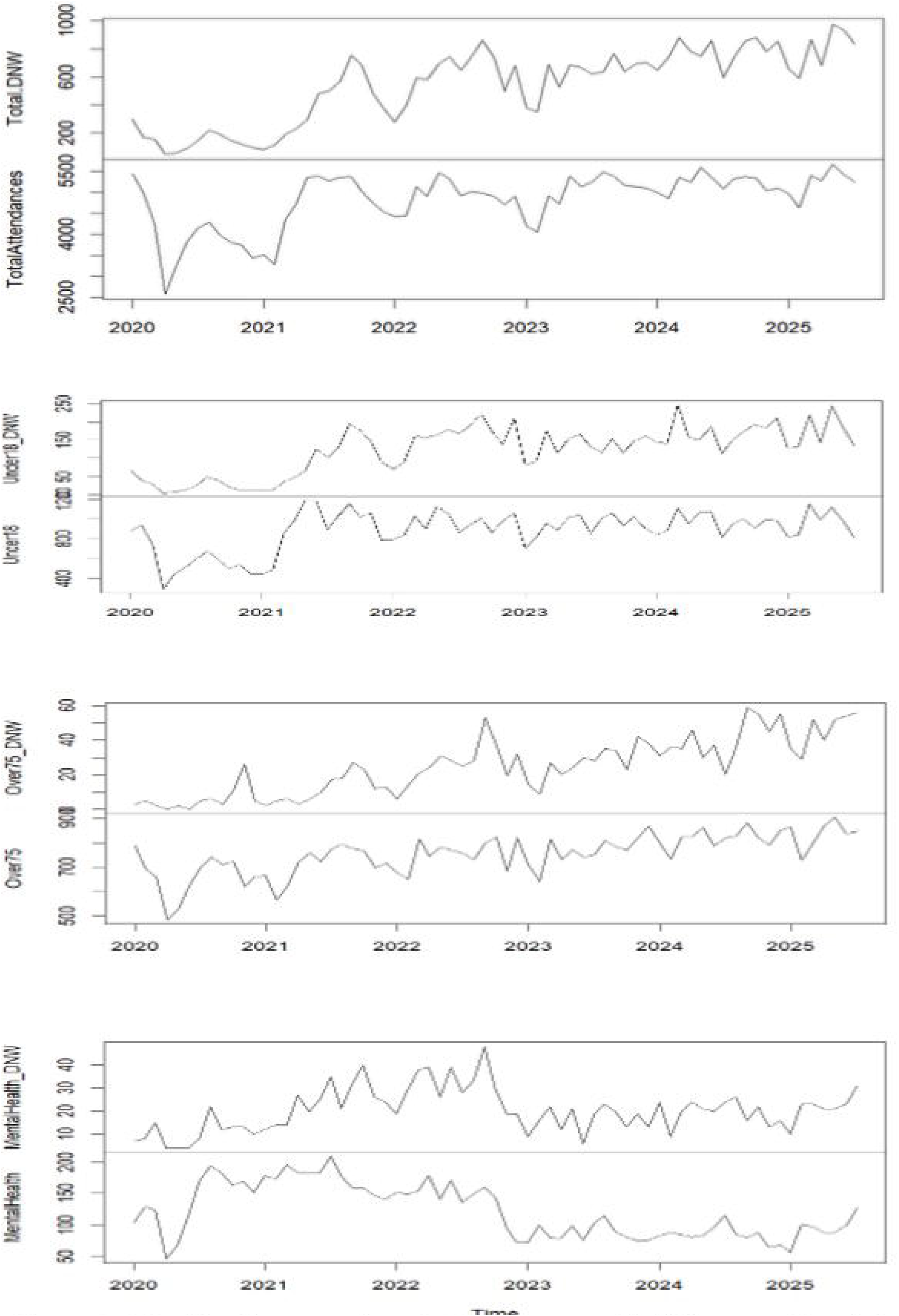
Showing the proportion of attendances and DNW across the whole population and each of the vulnerable groups.

**Figure 4.**
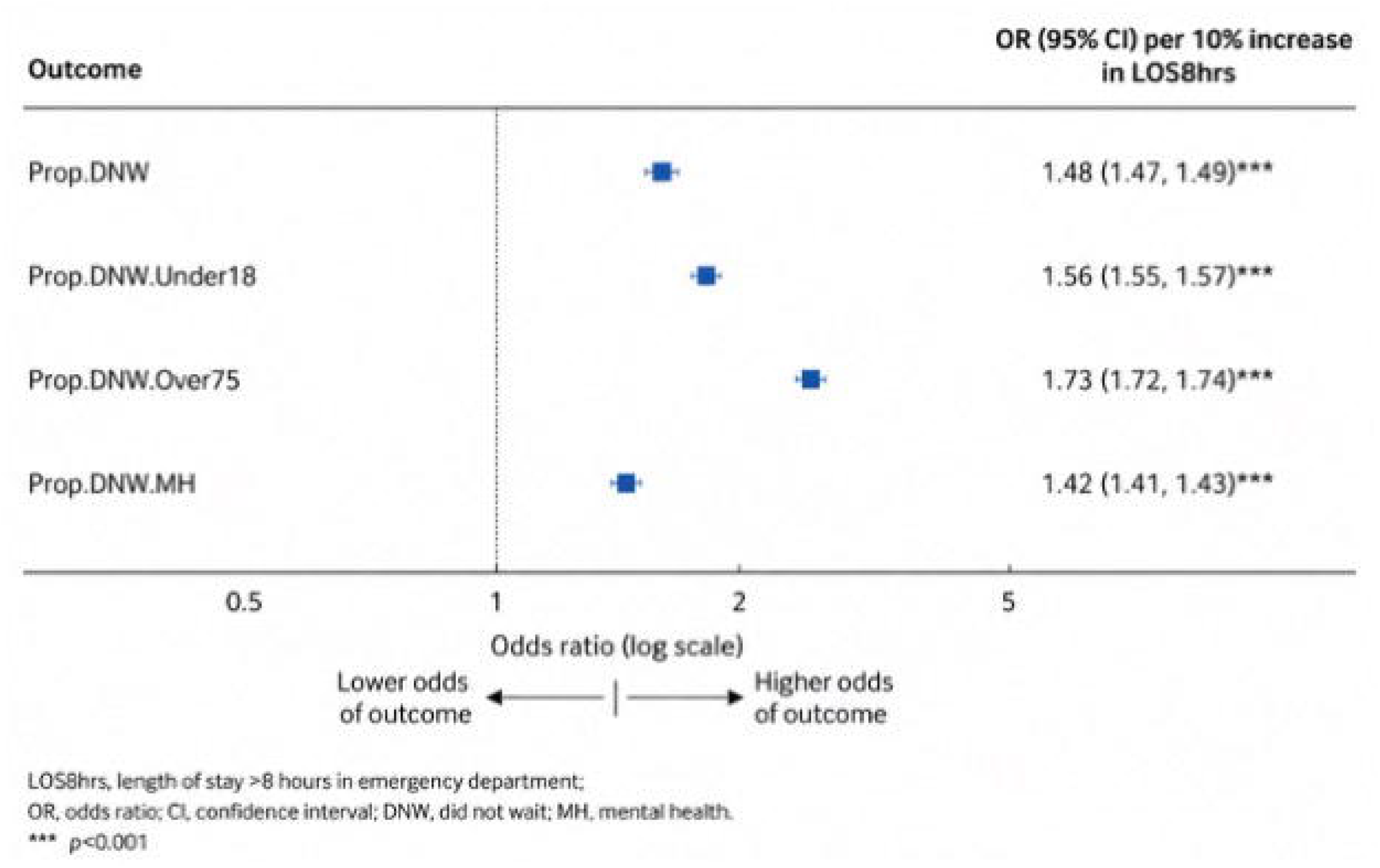
Showing generalised estimation equation model with binary outcome, used to model the proportion of those who did not wait (Prop. DNW) depending on the percentage of those who waited more than 8 hrs (LOS8hrs)

## Discussion

From Figure 2 there it is clear that declining 4-hour compliance is associated with rising length of stay, particularly lengths of over 8-hours. This suggests that long lengths of stay are not random or isolated phenomena, but are tightly linked to overall hospital flow performance. The association with LOS>8h implies that early system congestion rapidly propagates into prolonged delays, influenced by additional downstream constraints such as bed availability and discharge delays. This figure suggests that interventions undertaken to ease pressure and overcrowding within the ED and hospital site have made little impact on reversing the downward trend.

The association between the proportions of those that DNW in the individual vulnerability groups and the LOS>8h within the ED revealed interesting patterns. A lower proportion of patients with mental health related attendances DNW compared to the general population (42%, compared with 48%) suggests that patients with mental health attendances were less affected than the general population when LOS>8h, a finding which was unexpected. This could be due to triage pathways protecting this vulnerable patient group. However, as the group with the highest proportion of DNW generally (from 5% to 25%), the numbers of this patient group discharging without being seen remain significant.

The data clearly shows that the over 75-year group are more affected when LOS>8h increases. This could be due to a multitude of factors, including co-morbidities and frailty which make waiting in a crowded department more difficult, or perhaps a greater generational social responsibility in this age group, meaning that they choose to leave without being seen as they have decided that their need is not as great as others. As a group that typically does not DNW, even increases in the odds of this group leaving without being seen raise the population risk of DNW in this group from around 1% to 6%. Regardless, it is concerning that this age group, which are typically more frail and co-morbid are being disproportionately affected by crowding and exit block within the ED. Paediatric patients who DNW also showed a significant increase in DNW when LOS>8h increases, from 5% to 20%. Like the >75 group they are also affected by factors which make waiting more difficult when the ED is crowded, such as space to play and access to food and areas to rest. Initiatives to target more rapid assessment for these vulnerable groups need to be considered but would need additional resource to avoid prejudicing the care of other patient groups as an unintended consequence.

Despite the valuable insights provided by this study, several limitations should be considered. First, the use of age >75 as a proxy for frailty may not fully capture the complex nature of frailty in the ED population. Secondly, information bias may have occurred if the recorded variables were incomplete or inaccurately captured on our IT systems, however, this is less likely to apply to age and more likely to apply to recording of patients GP details or presenting complaint.

This study may be subject to bias. Miscalculation bias could arise due to the use of age >75 as a proxy for frailty. Information bias might result from incomplete or inaccurate data capture, especially for variables like GP registration status or presenting complaint. As groups of patients were compared proportionately against their attendance group, this reduced the risk of baseline bias, ensuring that differences in the monthly attendances of each vulnerable group were adjusted for.

This data is complex, but the results are largely unsurprising. The busier the department, the more patients that choose not to wait. Where this data causes concern is that it shows that as LOS>8h increases, not all patient groups are affected proportionately, with paediatric and elderly populations being more affected that the general population. With Emergency Departments being the last resort point of care access for many patients it is clear that overcrowding and exit block is driving inequality of care in certain vulnerable groups of patients. While the findings of this study provide valuable insights into the relationship between emergency department overcrowding and vulnerable patient groups, the results may be most generalisable to similar hospitals with high patient volumes and similar crowding issues. This is a healthcare emergency and consideration needs to be given at local and National level to solutions to improve emergency care access across all patient groups.

This study received no external funding.

## Data Availability

All data produced in the present work are contained in the manuscript

## Key messages statement

### What is already known on this topic

Emergency department (ED) overcrowding has been widely recognized as a factor contributing to delays in patient care, increased mortality, and worsened health outcomes. The Royal College of Emergency Medicine has identified crowding as a key issue influencing ED performance, but research on how specific vulnerable patient groups are impacted by long waits is limited.

### What this study adds

The findings demonstrate a clear association between lengths of stay of >8h in the ED and increased DNW rates, this is strongest among elderly and paediatric patients. These results provide new evidence on how overcrowding exacerbates inequalities in access to care for vulnerable patient groups.

### How this study may affect research, practice or policy

This study highlights the urgent need for targeted interventions to mitigate the effects of overcrowding on vulnerable patient populations. The findings could inform future research on the broader impacts of overcrowding, prompting further studies to explore the specific needs of vulnerable patients.

